# Understanding digital health technology implementation in rehabilitation: Development of the Rehabilitation Technologies Implementation model

**DOI:** 10.1101/2025.07.11.25331266

**Authors:** R.C. Stockley, H. Gooch, K. Jarvis, C. Watkins

## Abstract

**Background:** Technologies comprise a broad range of applications with the potential to transform outcomes for the millions of people requiring rehabilitation each year. Despite this potential, many technologies are not successfully implemented into clinical practice. No single consistent approach or model is used to support their implementation into any form of rehabilitation. This study aimed to understand the influences upon rehabilitation technology implementation in the UK’s National Health Service and, using this understanding, develop a comprehensive implementation model to support the adoption of rehabilitation technologies.

**Methods:** A multi-methodological approach comprising qualitative enquiry and literature reviews was used to identify, analyse and group key factors that influenced rehabilitation technology implementation.

**Findings:** Seven themes, identified from qualitative enquiry of 48 participants, representing 8 stakeholder groups in the UK, were integrated with published technology and implementation models to generate five key domains that comprise Rehabilitation Technology Implementation Model (RiTe). These domains were: evidence, technology, users, team and organisation.

**Interpretation:** The RiTe model provides a novel and comprehensive understanding of the key factors that influence rehabilitation technology implementation and can be used to plan, support and evaluate implementation efforts. The demands of rehabilitation require repeated, frequent and prolonged participation in interventions by users with variable needs, which means that barriers and facilitators are likely to exert a magnified effect upon the process and success of implementation. Consequently, the key influencers of rehabilitation technology implementation identified in this study provide a critical opportunity to understand factors which could influence technology uptake in other clinical specialties, attesting to the value of the RiTe model to both rehabilitation and wider healthcare.

## Background

Technologies have significant potential to transform healthcare^1^ and particularly to benefit the 2.4 billion people globally who require rehabilitation after illness, injury or disease.^2^ Rehabilitation technologies can be used to deliver, support, or augment aspects of rehabilitation and can overcome geographical challenges to access care, promote self-management, deliver more effective, or a greater intensity of, treatments, and motivate individuals to participate in regular training. There are a broad range of rehabilitation DHTs including remote consultations (telerehabilitation), virtual reality, gaming, sensors, applications (apps), robotics, exoskeletons and muscle and nerve stimulators.^3^ These DHTs form part of a rapidly growing health technology market; currently worth around US$316 million.^4^

Rehabilitation is provided by healthcare professionals across hospital and community settings and typically has more than one user group (staff, service-users and carers). Rehabilitation requires service-users, often having a range of cognitive and physical problems, to engage frequently, with technologies for weeks or months to gain benefit.^5,6^ The range of settings, needs of different user groups and the sequalae of conditions for which rehabilitation is sought, may go some way to explaining why, despite compelling evidence of the benefits of many forms of rehabilitation technologies,^7–11^ the implementation of rehabilitation DHTs into clinical practice remains suboptimal and highly variable.^12–15^

However, despite these challenges in implementing DHTs into rehabilitation, there are no comprehensive models that guide rehabilitation DHT adoption.^16^ Implementation models provide useful strategies to guide adoption of evidence-based interventions and innovations into routine practice, increasing the chances of successful and long-term adoption which benefits care.^17^ Consequently, this study aimed to generate a comprehensive model to support implementation of rehabilitation DHTs into clinical practice. This model and associated outputs will not only have benefit to the millions of people who could utilise DHTs within rehabilitation but will also provide important insights into technology implementation with direct relevance across healthcare.

## Methods

A multi-faceted methodology was utilised to reflect the complexities of implementing DHTs. This comprised qualitative exploration of stakeholder experiences into a national healthcare system (UK’s National Health Service), and three reviews of research evidence, detailed below. These findings were integrated to synthesise learning from practice with research and produce a comprehensive model of rehabilitation DHT implementation.

### Qualitative exploration

#### Identification of stakeholders

Stakeholder analysis identified key people for the qualitative interviews. Stakeholders were defined as people or groups who have influence or interest in DHT implementation in rehabilitation services, or who may be impacted by the outcomes of DHT implementation.^18,19^

#### Qualitative interviews

Opportunities to participate in the study was advertised via professional and social networks. Individuals representing NHS sites submitted expressions of interest, detailing their service, clinical area and the characteristics of their populations.

Sites were purposively selected to represent a mixture of urban and rural locales, deprivation levels and ethnicities.^20,21^ Within each site, exponential discriminative snowball sampling (via email) was used to recruit individual participants with diverse personal characteristics. This strategy also ensured representation of all identified stakeholder groups, across a range of clinical speciality areas.^22^ Individuals who wished to participate contacted the research team and were not previously known to the research team member who interviewed them.

Service-users and their carers were recruited separately from patient networks to ensure that they felt comfortable to give their views without concerns of potential influences on current or future care. Stakeholders who developed or regulated new technologies were recruited from professional and social networks.

After providing informed consent, participants self-reported their age, gender, ethnicity, current occupation and prior level of technology use.^23^ They completed a remote semi-structured interview using Microsoft teams with one of three (all female) members of the research team who are all both experienced therapists and interviewers (RCS, HG, KJ).^24^ Interviews lasted less than one hour.

The interview schedule was based upon normalisation process theory to enable exploration of the DHT-based intervention’s meaning (coherence), beliefs about it (cognitive participation), actions needed for its implementation (collective action), and evaluation of its implementation (reflexive monitoring).^25^

#### Analysis

Interviews were transcribed verbatim, checked for accuracy and inductively coded by each interviewer using NVivo.^26^ Field notes were used to add context to interview findings. Similar codes were grouped into themes. A constant comparison approach to analysis enabled iterative development and refinement of codes and themes. The first two interviews in each stakeholder group were coded by pairs of researchers establishing consistency in approach and interpretation.^27^ Data saturation was achieved when no new codes or themes were generated.^28^

Ethical approval was provided by the UK Health Research Authority and the University of Central Lancashire’s Health research ethics committee (319785, 0425,01086).

### Literature reviews

Three linked literature reviews were undertaken and comprised: (i) an integrative systematic review of models, theories and frameworks used in reports of rehabilitation DHT implementation for people with neurological conditions,^29^ (ii) a scoping review of the behaviour change approaches used in digitally supported stroke rehabilitation,^30^ (iii) a hermeneutic literature review to identify and critically consider models theories and frameworks of technology adoption across healthcare relevant to rehabilitation, had been used in more than one healthcare setting, or to implement or evaluate more than one form of technology.^31^

### Model generation

Themes generated from analyses of the qualitative interviews were used to generate an initial model. The components of the models, theories and frameworks (herein collectively called models) from across healthcare and within neurological rehabilitation identified in the integrative and hermeneutic systematic reviews, and the behaviour change approaches identified in the scoping review, were triangulated with these themes. If a component of a model or behaviour change approach did not clearly align to a generated theme, interview codes were revisited to check if any reflected the construct presented by the component of the model or behaviour change approach. If there were no codes that adequately represented the component, it was added as a subsection of the most appropriate theme. The resultant themes were then grouped into overarching domains.

## Results

### Qualitative exploration

#### Identification of Stakeholders

Key stakeholders comprised: adults who are using, or have used, digitally supported rehabilitation (service-users) and carers, clinicians who have adopted, attempted to adopt, or are using DHT for rehabilitation, those responsible for an organisation’s information technology (IT), people in organisations leading innovation (including service improvement, commercialisation and digital project leaders), organisational gatekeepers (including service leaders, mid-level or senior management), organisation regulatory leads (including information governance, or clinical safety personnel) and people developing, or regulating the use of new technologies (DHT companies, regional and national innovation and standards agencies).

#### Qualitative interviews

Ten NHS sites in England, Scotland and Wales were selected. Sites demonstrated a range of indices of multiple deprivation (IMDDs), a mix of rural and urban communities, and served varying levels of estimated ethnic minority service-users (see Table 1).

**Table 1.** Location and service-user characteristics from sites in the qualitative interview study.

| Site Location | Estimated<sup>#</sup> |  | IMD (rank in nation)* |
| --- | --- | --- | --- |
|  | Service-users from an ethnic group (%) | Rural and/or Urban locale |  |
| NorthWest | ≤10 | Mix | 1 |
| Wales | ≤25 | Urban | W13 |
| Greater London | ≤50 | Urban | 146 |
| NorthWest | ≤10 | Urban | 4 |
| NorthWest | ≤25 | Urban | 10 |
| Central Scotland | ≤25 | Mix | S8 |
| NorthWest | ≤50 | Mix | 14 |
| SouthWest | ≤25 | Mix | 88 |
| NorthWest | ≤10 | Mix | 49 |
| Wales | ≤10 | Mix | W14 |

<sup>#</sup>estimated by participating staff at these sites \*Rank for each site within their nation based on clinical commissioning group locations (England), mean average of regions covered by Health Boards in Scotland and percentage of most deprived 10% Lower Layer Super Output Area (LSOA) in Wales. <sup>20,21,32</sup> 1=most deprived. S denotes Scottish IMD Rank, W denotes Welsh Rank IMD for Health Boards.

Interviews were conducted with 48 participants (52% female) over 13 months (April 2023-May 2024). Participants were a median age of 50-59 years (range: 18-69; n=46; 2 missing) and of largely white ethnicity (White British: 44, Asian: 3, Other Ethnic group:1). All participants completed the interviews on their own, their roles and demographics are shown in Table 2.

**Table 2.** Characteristics of interview participants.

| Participant group | N | Gender (F:M) | Median age range years, (range) |
| --- | --- | --- | --- |
| Service-users | 8 | 3:5 | 60-69(50-69) |
| Digital developers | 3 | 0:3 | 30-39(30-49) |
| Clinical entrepreneurs* | 2 | 1:1 | 40-49<sup>#</sup> |
| Innovation leads | 3 | 1:2 | 50-59(40-59) |
| Staff involved in rehabilitation including | 21 | 14:7 | 50-59(18-69) |
| - Rehabilitation medicine | 2 |  |  |
| - Allied health professionals | 14 |  |  |
| - Nursing | 1 |  |  |
| - Support staff | 4 |  |  |
| Service level managers | 5 | 4:1 | 40-49(30-59) |
| Organisational leaders | 2 | 1:1 | 40-49<sup>#</sup> |
| Regulatory leads (including IT and IG) | 4 | 1:3 | 40-49(30-69) |

\*Clinical entrepreneurs had dual roles as a clinician (n=2) or manager (n=1). <sup>#</sup> median age range only reported to ensure participants cannot be identified.

Participants demonstrated experience of using a range of rehabilitation DHTs for several conditions (Figure 1) Most participants (n=44) predominantly based their answers upon their experiences of one form of DHT. The remainder based their answers upon a range of DHTs (n=4). From those that based their answers upon one technology, they were used in the community (n=16), in hospital (n=21) or across both community and hospital (n=7).

**Figure 1.**
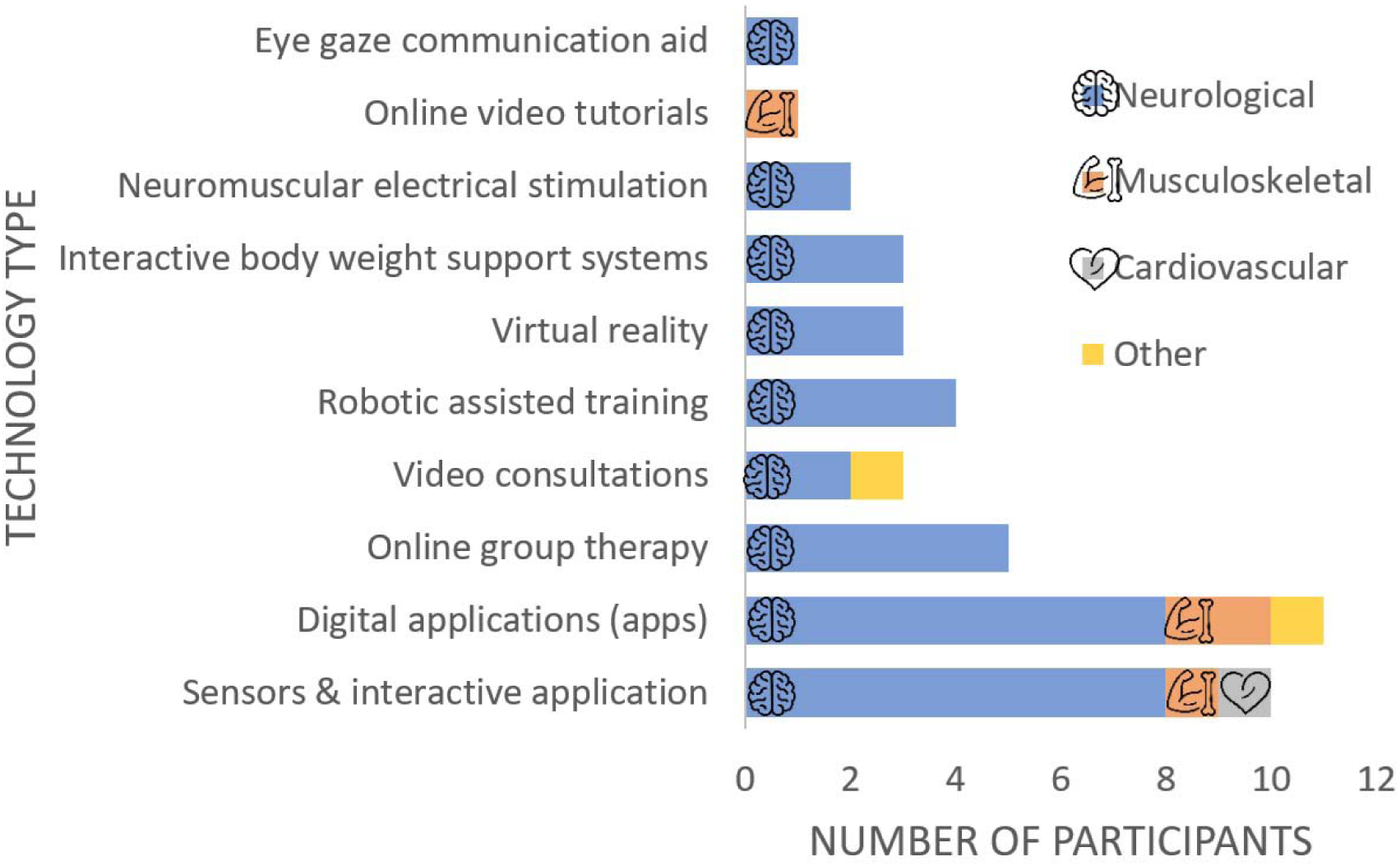
Bar chart to show clinical area and technology type reported by interview participants. Other= management of long-COVID (n=1) and general wellbeing and sleep (n=1)

Interviews generated 1308 codes which were grouped into themes (see coding tree and example quote in supplementary material).

Codes coalesced around 7 themes:

1. a description of the technology and its features, 2. the perceived need for the technology, including the potential benefit the technology was anticipated to deliver to services and service-users, 3. the key roles involved in the implementation process, 4. navigating the processes and procedures to implement the technology, 5. collaborations and communication needed between different individuals and groups within and outside the organisation, 6. key factors that influenced the implementation of the technology, and 7. understanding the response to DHT implementation both locally and more widely. The views and needs of different user groups were integral across several themes.

### Literature reviews

Findings of the review of the implementation models and behaviour change approaches are published elsewhere.^29,30^ In brief, 11 implementation models^33–43^ had been used to implement or evaluate DHTs used in neurological rehabilitation. None were specific to rehabilitation technologies and only two were explicitly developed for technology adoption.^33,44^ The components of all 11 models aligned to the themes generated from interviews and spanned five areas, (i) factors affecting individual’s ability and willingness to engage with DHTs (ii) user experience of DHTs; (iii) the content of the intervention supported or provided by DHTs; (iv) access to DHTs and (v) methods and mechanisms of supporting use.^29^

Few studies overtly articulated any behaviour change approaches utilised in rehabilitation DHT interventions: those studies articulating any form of behaviour change, reward (and threat), feedback and goal setting were most common. These reinforced previously identified themes exploring the suitability of DHTs for the user and promotion of user engagement.^30^

The hermeneutic review^31^ identified 6006 studies that had developed, used, or evaluated models (theories or frameworks) for technology adoption. Nineteen models, that had been used in more than one healthcare setting, with more than one health condition, and that articulated generic principles of technology adoption, were selected to triangulate with the other literature reviews and interview study findings to inform the model.^45–65^ From these, 7 models were not intentionally developed to implement DHTs but had been used to understand, plan, or evaluate technology adoption,^56–58,61–63,65^ whilst 12 were primarily developed to support or evaluate technology adoption in healthcare settings. ^44–47,50–54,59,60,64,66^

### Synthesis

Components of the models found in the three literature reviews were mapped to the interview’s seven themes, and largely reflected the components of the models identified in the literature reviews, reinforcing the credibility of the interview findings. However, there were no themes from the qualitative interviews directly related to objective evaluation of the implementation which necessitated combining findings from models identified in the literature reviews with the theme of ‘understanding the response to DHT implementation’. From this mapping process, the five overarching domains that comprise the RiTe model were generated (Figure 2, Table 3).

**Figure 2.**
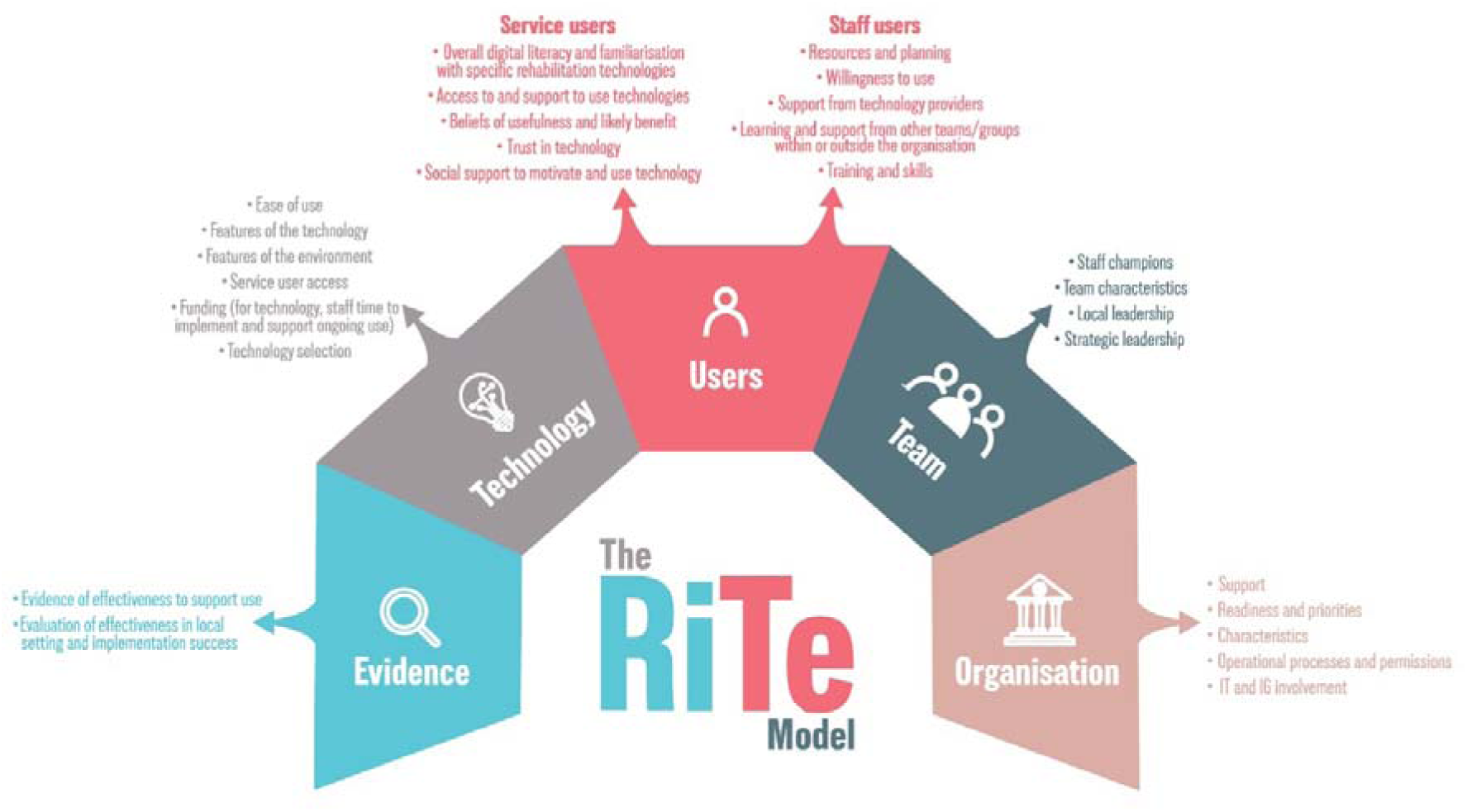
The Rehabilitation Technology (RiTe) Implementation Model

**Table 3.** Summary of the model domains.

| Domain | Definition | Constructs | Construct definition |
| --- | --- | --- | --- |
| Evidence | Using and collecting evidence of the effectiveness of the technology and its implementation | Evidence | Evaluating the available evidence to inform technology selection and to evaluate its use in the service into which it is implemented. Key features include: considering the applicability, independence of, strength and quality of evidence evaluating the effects of the technology to the intended setting. |
|  |  | Evaluation | Identification and rigorous measurement of the key outcomes that can be used to judge the success of the implementation process and the effectiveness of the technology. This includes collecting evidence of service users outcomes, service benefits, success of the implementation and methods to identify any unintended consequences. |
| Technology | The technology being implemented | Ease of use | The physical, technical and simplicity of initially setting up, and then using the technology regularly. Features to consider include the time take to set up, requirements for logging in (time and frequency), the perceptions of the ease of use and user expectations, the familiarity (or similarly to other tech) of the technology to user groups, clarity of the user interface enabling it to be accessible and easily navigable |
|  |  | Features of the technology | The suitability of the technology for its intended use.<br>Features include the ability of the technology to address the target of rehabilitation, its practicality to be used in appropriate settings, its clarity to use, its ability to be adapted and personalised so it is optimally suitable for patients, provision of feedback/data for all users, facilitates service user engagement (by supporting independent use, facilitating self efficacy, providing reminders, prompts etc to continue engagement, providing novelty and fun, motivating experience) |
|  |  | Features of the environment | Consideration of the intended environment the technology is to be used in. This includes physical factors such as space for equipment but also connectivity, hardware etc required. For a home setting, pertinent factors include: convenience, comfort, space. For clinical setting factors include: accessibility, space and additional equipment needed, portability, infection control considerations |
|  |  | Factors influencing service user access | Factors that influence who can access and utilise the technology. Factors relate to digital inclusion. |
|  |  | Funding (tech and staff)* pre implementation | The process of identifying funding and the processes to utilise funding to purchase the technology and maintain it. Includes understanding initial and ongoing costs, procurement processes and consideration of the time for implementation, ongoing training and contingencies within funding to react to unforeseen events. |
|  |  | Selecting the 'right' technology * pre-implementation | Understanding the available technologies on the market and their selection. These include ways of finding the available technologies, the opportunity to trial a potential technology in the clinical setting and the research evidence supporting its effectiveness (see Evidence and Evaluation domain). |
| Organisation | The organisation within the which the technology will be implemented. There may be multiple organisations e.g. the hospital system, a regional commissioning group/organisation, a national healthcare system (e.g. the NHS) | Support | Sources of support for technology implementation that exist within the organisations. These may include formal (e.g. innovation teams) or informal (e.g. experienced colleagues). |
|  |  | Readiness and priorities | The current ability and willingness of the organisations to implement technology. Features include familiarity with technology implementation, alignment with other organisation level changes (which maybe either complementary or conflicting), alignment with local or national priorities. |
|  |  | Characteristics | The attitude towards, and previous experience of, implementing (technological) change within the organisations. This includes previous positive or negative experiences of innovation, willingness to embrace change, effective channels of communication between different stakeholders within the organisation (e.g. IT, Clinician team, IG). |
|  |  | Operational processes and permissions | Understanding of the operational processes and permissions that must be completed to implement a new technology. This includes clear guidance of the requirements for, and capacity to undertake, timely assessment of data protection, infection risk, procurement processes and other risk assessments (depending on the equipment). |
|  |  | IT and IG involvement | The Information technology and governance scrutiny required before implementation of a new technology or addition of new features to an existing technology. This comprises: alignment of the technology to be implemented to local and national IT and IG policies, the information required to make judgements of IT and IG implications and the availability and accessibility of IT and IG teams to provide timely advice, support and undertake appropriate assessments. |
| Team | The values, beliefs and performance of the team that is implementing or will be using the technology, and alignment of the technology to be implemented with other team priorities. | Staff champions | An identified individual or group from within the team who will lead and support the technology implementation. These individuals are recognised to be respected and embedded within the team, are motivated and enthusiastic about the technology to be implemented and demonstrate resilience to setbacks, problem solving skills, a growth mindset and have interpersonal skills that enable them to be are facilitatory and encouraging to others. |
|  |  | Team characteristics | The culture, beliefs and behaviours of the team who will be using the implemented technology. Key influences include the presence of a shared vision for the benefits the technology can deliver, clear and strong channels of communication within the team, a supportive team ethos, embedded mechanisms to include the experiences and views of the service users, a culture of problem solving and their physical proximity to each other and wider stakeholders within the organisation. |
|  |  | Local leadership. This is local manager or team leader who has oversight for the day-to-day running of the team/service. | Commitment to support the time, processes and change needed to implement and continue to use the technology. Key features include: knowledge of internal processes and networks, flexibility and authority to support the time needed to implement the technology, understanding of change management. |
|  |  | Strategic leadership<br>This is an organisational leader who has insight and (some) influence upon the strategic direction and priorities of the service and wider organisation. | Support for the technology implementation at a strategic level within the organisation. Key features include: championing of project with senior leadership/management, provision of overt support for the technology adoption, alignment of technology implementation to strategic priorities for the organisation and nationally, financial support for funding (to purchase and maintain technology and to enable staff time to implement). Without this support, implementation is likely to fail. |
| Users | The end users of the technology, comprising service users (patients) those that care for them and staff | Service users | The beliefs, experiences and needs of service users (and those that care for them) to access and utilise the technology.<br>Factors include: digital literacy (familiarity with technologies in wider life as well as with healthcare technologies), support to access and use the technology provided by the clinical service, their beliefs of the usefulness, trust, and evidence of benefit that the technology can deliver, presence of social support to motivate and engage. |
|  |  | Staff users | The beliefs, experiences and needs of staff users to use the technology as part of their usual practice. |
|  |  | - Resources and planning | Recognition that implementation of technology requires obvious (e.g. funding) and hidden resources (e.g. time) and technology implementation will change models of current service provision. |
|  |  | - Willingness to use | Understanding of the evidence underpinning a technology, beliefs and values of the benefit offered by the technology, plus staff users confidence to implement and use the technology. This includes staff interest in technology and their current ability to embrace change to their daily practice. |
|  |  | - Support from technology providers | The provision of timely, tailored support from technology providers to enable implementation, continued use, training and troubleshooting. Features include building relationships with an individual at the company, their physical presence on site when needed, availability for ongoing support particularly at ‘pinch points’ including ensuring compliance with IG, initial training and set up, managing glitches, updates, and provision of ongoing training. |
|  |  | - Learning and support from other teams/groups within or outside the organisation | Use of learning from others who have implemented or used the technology. This may guide, inspire, advises and can provide a community of practice |
|  |  | - Training and skills | The training and skills required so that staff are competent and confident to utilise specific technology and to support others to use it. Training may comprise informal or formal peer support and training undertaken in ring-fenced time. |

#### Evidence

This domain addresses the importance of using evidence to inform selection of the DHT and collecting evidence to evaluate the DHT’s effectiveness and its implementation. Research evidence informs technology selection which, depending upon its independence, strength and quality, will influence judgements of the technology’s applicability to the intended setting and its likely benefits. After implementation, evidence generated from clinical outcomes enables evaluation of clinical and cost-effectiveness, and identification and monitoring of unintended outcomes (for example, systematic exclusion of groups based on socio-economic or demographic factors).

#### Technology

This domain highlights that, unsurprisingly, the features of a specific technology influences both the choice to, and processes of, implementation. Key factors that influence the selection of a DHT include its suitability to meet clinical needs, consideration of costs versus benefits, initial and ongoing funding, and availability to trial and eventually procure the technology in the health organisation, in addition to evidence (as described in the ‘Evidence’ domain above). Critical consideration of suitability for the intended service-users, including how accessible and easy it would be, the ability of the technology to effectively and efficiently provide a desired targeted treatment as part of a novel, engaging rehabilitation intervention, the environment in which the technology will be used (home, community or hospital), and factors which may precipitate digital exclusion (for example, reliance on internet connectivity, large data demands) influence both choice of a technology and its implementation’s reach.

#### Users

This domain focuses upon the end-users of the technology and was divided into service-users (and their carers) and staff-users. Familiarity with technology, previous experiences, evidence and beliefs about the benefits and limits of technology influence the willingness and ability to use technology for both user-groups. The need for support from others was also highlighted, although by whom and how this was provided differed for the two user-groups.

Service-users value real-time, practical support to use specific technology and their uptake of technology is influenced by their familiarity with technology and digital literacy. The potential for exclusion of some service-users who do not have access to hardware or connectivity from digitally-based interventions was also recognised. The interviews indicated that support for service-users was largely provided by clinical staff (e.g. telephone instructions to support a service-user to log into the technology) with social support from family and friends motivating the technology’s continued use.

The need for staff-users to feel confident and competent appears instrumental to their use, or non-use, of technology. Confidence and competence could be achieved via several routes; staff-users value support from those who developed or provided the technology, particularly at ‘pinch points’ including initial training and set-up, managing glitches, technology updates, and provision of ongoing training. They also value support from colleagues in other organisations who had experience of implementation and use of similar DHTs, peer support from colleagues and more formal, ongoing training.

#### Team

This domain incorporates the values, beliefs and performance of the local organisational team that is implementing and using the technology. It highlights the importance of understanding the technology’s alignment with team priorities. The team’s characteristics, notably their commitment to a shared vision for the benefits of the technology, culture and ways of working influence implementation. Embedded, well-respected individual staff members given time and resource to ‘champion’ the technology adds legitimacy to the implementation process and appears pivotal in supporting other staff-users. Ongoing support from local leadership (team or department-level with responsibility for day-to-day team performance) and strategic leadership (senior management level) is critical to implementation, with the absence either of these roles potentially precipitating failed implementation. Local leadership is particularly important to facilitate ring-fenced time and resource for staff champions and local leads, supporting training activities and navigating the operational processes and procedures. The support of leaders who have insight into, and could influence, the wider strategic direction of the service and organisation provides a conduit to gain top-down support for the implementation, potentially removing some barriers to implementation and generating financial support for DHT purchase and maintenance.

#### Organisation

The organisation domain explicitly recognises the impact of the complex adaptive nature of healthcare organisations upon rehabilitation DHT adoption. Understanding and alignment of the technology implementation with national and local organisational policies and procedures positively influences adoption, along with the agility of the organisation to respond to and manage change.

Some of these processes are vital, nationally-mandated, non-negotiable and sometimes anticipated (for example, Data Protection and Information Assurance Assessment). Knowledge and understanding of the detailed requirements of both national and local processes (for example, procurement) positively influences the time and effort required for a DHT’s approval, purchase and implementation. Interview findings highlighted that the absence of this knowledge leads to frustration and can mean the abandonment of technology implementation altogether.

A culture of, and resources for, innovation are important organisational characteristics that influence the ease and success of timely technology implementation. Unsurprisingly, strong, visible and clear channels of communication between all stakeholder groups, most notably between those adopting the technologies into their service, significantly influences the implementation process. In particular, the involvement of the organisation’s IT and Information Governance (IG’s) team are key elements within the organisation domain, and their individual personnel’s availability, visibility and accessibility significantly influences implementation. Advice and guidance from the IT and IG teams is pivotal for local implementation leads to understand, fulfil, and gain approvals in a timely fashion. In turn, the alignment of the DHT with IT and other organisational priorities is an important influencer of the IT team’s prioritisation of local implementation efforts.

## Discussion

This study sought to explore the key influences upon DHT implementation in rehabilitation. It synthesised findings from qualitative research and literature reviews to provide a multi-faceted comprehensive understanding of the key factors, processes and people that influence technology adoption. This understanding was incorporated into the RiTe model. This model enables navigations of the often complex, confusing and opaque technology adoption process. Despite a plethora of technology implementation models and frameworks,^67–70^ to our knowledge there are no recognised models, theories or frameworks that have been explicitly developed to support DHT implementation in rehabilitation, underlining the impact of this work.^16^ The novelty of the RiTe model is further strengthened by its use of extant models of implementation from technology, wider healthcare and innovation in combination with real-world experiences gained from qualitative interviews. We believe this adds important specific context to the understanding of implementation of rehabilitation technology not fully captured by previous models. Whilst requiring empirical evaluation, the combination of qualitative, real-world data, reinforced by elements from other theories, frameworks and models of implementation, innovation and digital adoption within the domains of the RiTe model reinforces its validity.^12,16,71^

The five domains included in the RiTe model provide a multi-level understanding of the key factors influencing DHT implementation in rehabilitation, and accordingly, the model should ideally be used in its entirety. Whilst it is recognised that some elements of the RiTe model may seem distant or irrelevant to local technology implementation, in the qualitative interviews, participants reported that failure to appreciate all potential influences upon the process, including those which initially seem remote to the local implementation effort (such as changing organisational priorities) generated significant barriers to implementation. This importance of this holistic approach to technology adoption is supported by the multi-factorial reasons for failure of technology adoption reported in healthcare; these include the knowledge and views of both the service-user and the clinician,^72–78^ alignment between the clinical need and the technology,^12^ cost, access to and usability of the technologies^12,77,79–81^ and wider organisational factors.^82–85^

There are several limitations of this work. As participants volunteered to take part in interviews, it is likely that those that had been unsuccessful in their implementation efforts, although explicitly sought by the researchers, would be less likely to volunteer which may bias findings. Participants rarely mentioned evaluating the effects of the DHT or the implementation process. This may because many technology implementation projects were relatively recent and so the long-term spread or reach of the technology had not yet been investigated. Alternatively, this could indicate a lack of clarity of the specific effects of the implemented technology, an unfamiliarity with implementation models or a belief that the implementation process was complete once the technology was being used in the clinical setting.^86^ However, learning from previous implementation efforts, both successful and unsuccessful is important in DHTs as they continue to become more commonplace in healthcare, and prioritised in national and international strategies.^87^ This underlines the need for targeted tools to support implementation in clinical settings, including evaluation (such as those emerging from the RiTE model and provided here: https://advancingrehab.com/projects/dare/dare-phase-1/).

Whilst the lack of evaluation of the implementation process limited the qualitative research findings, the strength of the plural methodological approach we employed in this study enabled established models of technology adoption, and implementation, to be used to expand and consolidate this domain.

As the largest universal healthcare system and employer of skilled professionals gloablly,^88^ learning from the UK’s NHS provides a critical exemplar of the key factors that influence technology implementation both in rehabilitation and more widely. Although the qualitative interviews included NHS organisations in England, Scotland and Wales, the integration of globally developed and utilised implementation models (retrieved in the literature reviews) into the RiTe model supports its validity outside the UK, and in other healthcare systems. However, it is recognised that global healthcare systems (such as insurer funded or out-of-pocket services) may have different or additional factors to those included in the RiTe model (for example, the requirements of health insurers, challenges of geography and different national and local priorities). Consequently, future research should seek to establish the validity, test and adjust the RiTe model in range of settings to ensure it realises its potential to support timely and successful adoption of technologies into rehabilitation.

## Data Availability

De-identified summarised interview data (code book) and qualitative enquiry protocol are available from: 10.17030/uclan.data.00000482 These resources will be freely available immediately upon publication and until at least 5 years after publication to anyone.

https://uclandata.uclan.ac.uk/id/eprint/482

## Data sharing

De-identified summarised interview data (code book) and qualitative enquiry protocol are available from: <u>10.17030/uclan.data.00000482</u> These resources will be freely available immediately upon publication and until at least 5 years after publication to anyone. Protocols for each literature review are available from the full text publications (Doi: 10.1177/11786329241229917 and 10.2196/48725) and here: https://osf.io/6fdbj/

